# Bone morphogenetic protein 2 is a new molecular target linked to nonalcoholic fatty liver disease with potential value as non-invasive screening tool

**DOI:** 10.1101/2022.03.17.22272494

**Authors:** Patricia Marañón, Carlos Ernesto Fernández-García, Stephania C Isaza, Esther Rey, Rocío Gallego-Durán, Rocío Montero-Vallejo, Javier Rodríguez de Cía, Javier Ampuero, Manuel Romero-Gómez, Carmelo García-Monzón, Águeda González-Rodríguez

**Author notes:** **Correspondence**: Águeda González-Rodríguez., C/Maestro Vives 2, 28009 Madrid, Spain. These authors contributed equally to this work (Carlos Ernesto Fernández-García, Stephania C Isaza, Esther Rey). Instituto de Investigaciones Biomédicas Alberto Sols (Centro Mixto CSIC-UAM), 28029 Madrid, Spain.

## Abstract

Nonalcoholic fatty liver disease (NAFLD) is the commonest cause of chronic liver disease worldwide, being nonalcoholic steatohepatitis (NASH) its most clinically relevant form. Given the risks associated with taking a liver biopsy, the design of accurate non-invasive methods to identify NASH patients is of upmost importance. BMP2 plays a key role in metabolic homeostasis; however, little is known about its involvement in NAFLD onset and progression. This study aimed to elucidate the impact of BMP2 in NAFLD pathophysiology. Hepatic and circulating levels of BMP2 were quantified in serum and liver specimens from 115 biopsy-proven NAFLD patients and 75 subjects with histologically normal liver (NL). In addition, BMP2 content and release was determined in cultured human hepatocytes upon palmitic acid (PA) overload. We found that BMP2 expression was abnormally increased in livers from NAFLD patients than in subjects with NL and this was reflected in higher serum BMP2 levels. Notably, we observed that PA upregulated BMP2 expression and secretion by human hepatocytes. An algorithm based on serum BMP2 levels and clinically relevant variables to NAFLD showed an AUROC of 0.886 (95%CI, 0.83–0.94) to discriminate NASH. We used this algorithm to develop SAN (Screening Algorithm for NASH): a SAN < 0.2 implied a low risk and a SAN ≥ 0.6 indicated high risk of NASH diagnosis. This proof-of-concept study shows BMP2 as a new molecular target linked to NAFLD and introduces SAN as a simple and efficient algorithm to screen individuals at risk for NASH.

## INTRODUCTION

Nonalcoholic fatty liver disease (NAFLD), is becoming the most prevalent chronic liver disease worldwide and is considered the hepatic manifestation of the metabolic syndrome; consequently, it is highly linked with the most relevant features of this syndrome, as obesity, hypertension, dyslipidemia, central adiposity and insulin resistance or type 2 diabetes (T2D) ^1,2^. Indeed, the prevalence of NAFLD in obese and diabetic patients can be over 50%, whereas, the global prevalence of NAFLD is around 25% in adults, being higher in Western countries ^3,4^.

NAFLD encompasses a range of liver diseases from simple fatty liver (NAFL), mostly a benign non-progressive clinical entity defined as the presence of fat in > 5% of hepatocytes, to steatohepatitis (NASH), a more severe condition featured by steatosis, lobular and portal inflammation, and degeneration (ballooning) of hepatocytes, with or without fibrosis, which in turn can lead to more severe conditions of liver disease such as cirrhosis, portal hypertension and even hepatocellular carcinoma (HCC) ^5,6^. In this regard, while most of NAFLD patients are usually asymptomatic and only present a simple accumulation of fat in the hepatocyte (NAFL), a 44-59% of patients end up developing NASH with variable fibrosis stages. Up to 25% of patients with NAFLD progress to cirrhosis and approximately 7% to end-stage liver disease, needing for liver transplantation ^3^.

The prompt diagnosis and treatment of the early stages of NAFLD are important to prevent the progression to advanced stages of the liver disease. The determination of the lipid content of the liver can be an important challenge in terms of identification, treatment and control of the progression of the disease ^7^. Nowadays, there are no validated non-invasive diagnostic methods for quantifying NAFLD histological evolution, and liver biopsy is still the gold standard to differentiate fatty liver from NASH and/or fibrosis stages ^1^. However, liver biopsy is not recommended for routine use due to increased risk of bleeding and complications, and intra and interobserver variations. In the last years, many diagnostic non-invasive tools (mainly serological tests and imaging techniques) have been described, but they are not ready yet for widespread use, or to replace histological examination ^8,9^.

Bone morphogenetic proteins (BMPs) are secreted glycoproteins belonging to the transforming growth factor β (TGFβ) superfamily. More than 20 BMPs have been described so far and originally identified as growth factors involved in the differentiation of osteogenic cells, but it has been demonstrated that these proteins also play a critical role in the development of many cells types in various tissues, acting in cell proliferation and differentiation, tooth morphogenesis, organogenesis, embryonic development, apoptosis, chemotaxis and repair of a wide variety of tissues ^10,11^.

Regarding BMPs and liver physiology, these proteins are known to have a crucial role in different stages of hepatic development and also are implicated in adult liver homeostasis, as iron and glucose homeostasis ^12^. In this regard, while it has been demonstrated the pathophysiological role of some BMPs, such as BMP3B, BMP4, BMP6, BMP8B and BMP9, in animal models of NASH and NASH patients ^13-17^, little is known about the involvement of BMP2 in experimental models of NAFLD ^18^, and no data exist so far directly linking BMP2 to human NAFLD. Therefore, we sought to explore whether BMP2 could be related to NAFLD by assessing the expression of BMP2 in liver and serum from patients with biopsy-proven NAFLD as well as in an *in vitro* model of hepatic steatosis.

## MATERIALS AND METHODS

### Study population

This study comprises 115 patients with biopsy-proven NAFLD, 56 with simple steatosis (NAFL) and 59 with steatohepatitis (NASH) as well as 75 patients with histologically normal liver (NL) to whom a liver biopsy was performed during laparoscopic cholecystectomy. All patients were recruited from Hospital Universitario Santa Cristina (Madrid, Spain) and Hospital Universitario Virgen del Rocío (Sevilla, Spain). Inclusion criteria were: age between 18 and 75 years old, alcohol consumption lower than 20 g/day, no history of potentially hepatotoxic drugs and no analytical evidence of iron overload, autoimmunity and hepatitis B, hepatitis C and human immunodeficiency virus infection.

This study was performed in agreement with the Declaration of Helsinki, and with local and national laws. The Human Ethics Committee of the Hospital Universitario Santa Cristina (reference, PI-688A) and Hospital Universitario Virgen del Rocío (reference, C.I. 0359-N-15) approved the study procedures, and all participants signed an informed written consent before inclusion in the study.

### Clinical, biochemical and metabolic assessment

A complete clinical examination was performed to all patients studied. Body mass index (BMI) was calculated as weight (kilograms) divided by height (meters) squared. Blood samples of each participant were obtained, at the time of liver biopsy, to determine serum levels of liver enzymes, metabolic parameters and serologic tests using routine laboratory methods. Insulin resistance was calculated by the homeostasis model assessment (HOMA-IR) method ^19^.

### Histopathological assessment

Hematoxylin-eosin and Masson’s trichrome-stained paraffin-embedded liver biopsy sections showing more than 10 complete portal tracts were examined and interpreted by experienced hepatopathologists of each clinical site using the same internationally accepted criteria.

Briefly, steatosis was assessed as outlined by Brunt et al ^20^ grading percentage involvement by steatotic hepatocytes as follows: grade 0, < 5%; grade 1, 5-33%; grade 2, > 33-66%; and grade 3, > 66%. Simple steatosis or NAFL was defined as the presence of at least 5% of steatotic hepatocytes with or without mild lobular or portal inflammation but in the absence of features of hepatocellular injury (ballooning, apoptosis or necrosis) and fibrosis. On the other hand, minimal criteria for the histological diagnosis of NASH included the combined presence of grade 1 steatosis, hepatocellular injury and lobular inflammation with or without fibrosis. SAF score described by Bedossa *et al* ^21^ was used to categorize hepatic lesions of each liver biopsy studied and was considered as the gold standard method for diagnosis of NAFL and NASH. In addition, NAFLD activity score was also calculated to each liver biopsy according to Kleiner’s criteria ^22^.

### Cell culture and treatment

Human hepatoma cell line Huh7 (ATCC, Manassas, VA, USA) was maintained in Dulbecco’s modified Eagle’s medium (DMEM, Cytiva, USA) containing high glucose and antibiotics and supplemented with 10% fetal bovine serum (FBS) at 37°C with 5% CO2. For the steatogenic protocol, cells were treated overnight with palmitic acid (PA, Merk Millipore, Darmstadt, Germany) 750 nM in serum free DMEM with 1% BSA.

### Gene expression analysis

Total RNA from liver samples was extracted using the NucleoSpin RNA purification kit (Macherey-Nagel, GmbH & Co. KG, Dueren, Germany) following manufacturer’s instruction. For cell samples, plates were washed with phosphate buffered saline (PBS) and total RNA was extracted with TRIzol Reagent (Vitro, Sevilla, Spain). Reverse transcription was performed with ImProm-II™ Reverse transcription kit (Promega Inc., Madison, WI, USA) in a T100TM Thermal Cycler (BioRad Inc., Madrid, Spain). Quantitative Real time PCR (RTqPCR) was performed using Syber Green method and quantified with ΔΔCt method. All samples were run in duplicate and normalised by *36B4*.

Primer sequences used were: *BMP2*, 5’ CAACACTGTCGCAGCTTC 3’ (forward) and 5’ GAAGAATCTCCGGGTTGTTTTC 3’ (reverse); *CD36*, 5’ ATGTGTGTGGAGAGCGTCAACC 3’ (forward) and 5’ TGAGCAGAGTCTTCAGAGACAGCC 3’ (reverse); *36B4*, 5’ CAGGCGTCCTCGTGGAAGTGAC 3’ (forward) 5’ CCAGGTCGCCCTGTCTTCCCT 3’ (reverse).

### ELISA Assay

BMP2 concentration in serum samples from patients or supernatants from cell culture was measured using an ELISA kit (CSB-E04507h, Cusabio Technology LLC, Hubei, China) following manufacturer’s instructions. Absorbance from samples was interpolated to a standard curve using a four parameter logistic (4-PL) equation.

### Intracellular lipid content detection by Oil Red O

Cells were cultured in 12 mm coverslips and treated as described above. Then, cells were washed with PBS and stained with an Oil red O (ORO, Merk Millipore, Darmstadt, Germany) working solution (60% ORO/isopropanol w:v) and counter-stained with hematoxylin. Images were taken using an optical microscope Nikon Eclipse E400 (Nikon, Tokyo, Japan) equipped with a plan Apocromatic 20x objective (Nikon). Intensity of red stain was quantified using the FIJI software (NIH, Bethesda, MD).

### Statistical analysis

Kolmogorov-Smirnoff test was applied to evaluate if the variables were adjusted or not to a normal distribution. Qualitative variables are presented as relative frequencies. Quantitative variables are expressed as measures of central tendency (mean) and dispersion (standard deviation -SD-for patients’ data or standard error of mean -SEM-for experimental data). Data between groups were compared with Student’s t test for variables following a normal distribution and Mann-Whitney U test for continuous variables following a non-parametric distribution. Binary and multinomial logistic regression analysis adjusted for confounders were used to test the independence of the associations of dependent variable (NASH) with its significant correlates in univariate models and with those considered of clinical interest. In addition, multivariate regression models were constructed and parameters selected by stepwise method based on likelihood ratio test. Box-Tidwell procedure was used for testing linearity of logit and to obtain a linear logit the appropriate transformation of variables was used. The goodness of fit of the model was evaluated using the Hosmer-Lemeshow statistic and Pearson’s Chi-squared test as appropriated. In order to assess diagnostic accuracy of distinct regression models constructed, receiver operating characteristics (ROC) curves and area under the ROC curve (AUROC) were carried out. All statistical analyses were performed using the GraphPad Prism 6.0 software (GraphPad Software Inc., San Diego, CA, USA) and the IBM SPSS Statistics 24.0 (SPSS Inc., IBM, Armonk, NY) software with two-sided tests, with a p value of <0.05 considered as statistically significant.

## RESULTS

### Characteristics of the study population

Characteristics of all patients included in this study based on the histological diagnosis are shown in Table 1. Briefly, women were predominant in all the histological groups studied and NAFLD patients were significantly older than NL subjects. As expected, BMI was significantly higher in NASH patients compared to both NL and NAFL groups, and variables regarding glucose metabolism (glucose, insulin, HOMA-IR) and liver enzymes (GOT, GPT and GGT) were significantly different between all groups.

**Table 1.**
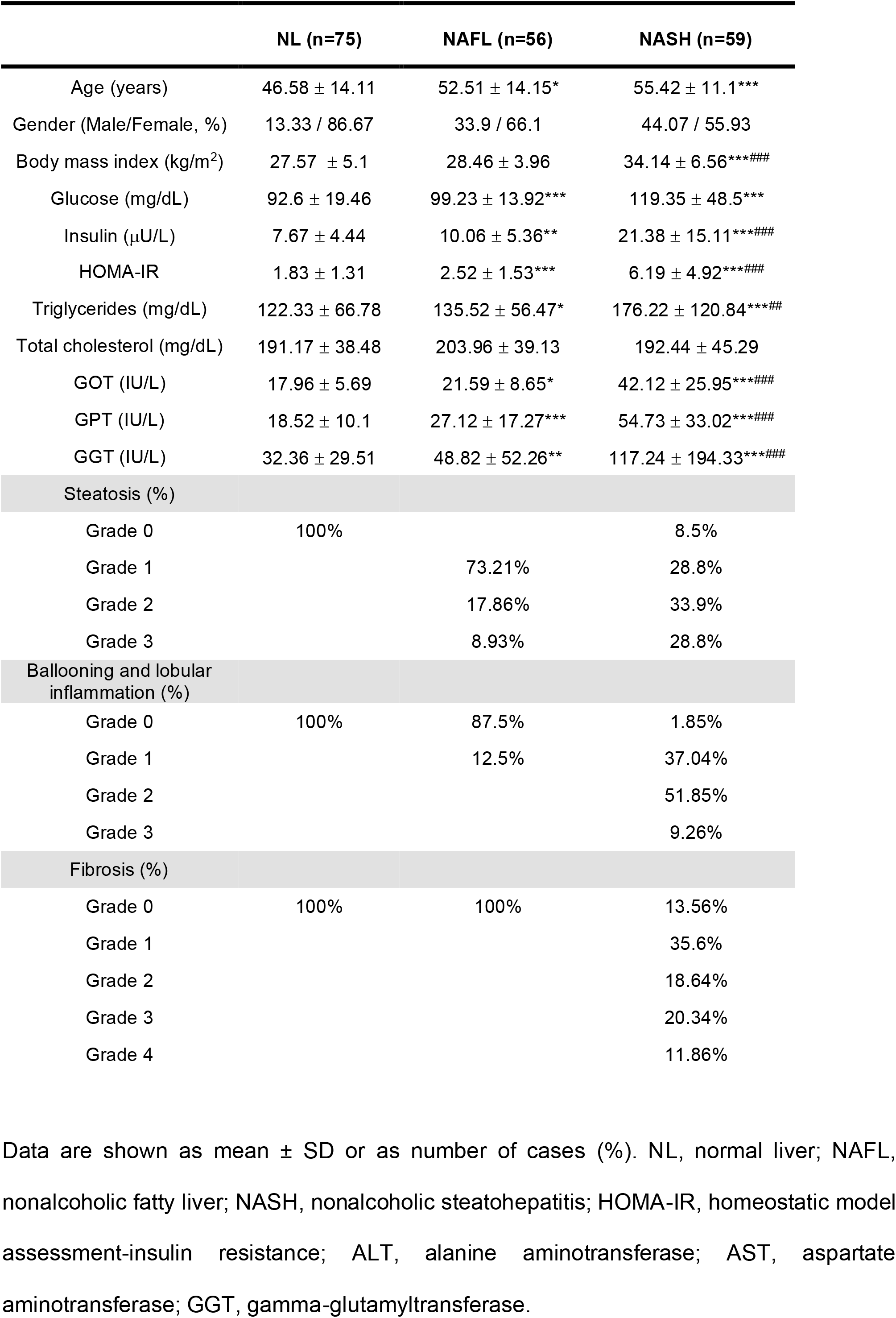
Characteristics of the study population.

### Hepatic BMP2 expression is increased in patients with NAFLD

We found that BMP2 gene expression was significantly increased in the liver tissue of NAFLD patients with respect to NL subjects (2-fold, p<0.0002; Figure 1A). Indeed, when compared with NL, hepatic mRNA levels of BMP2 were significantly higher in patients with NAFL (1.7-fold, p=0.0323) and NASH (2.6-fold, p<0.0001) (Figure 1B), in parallel with the increase of the liver injury. It was noteworthy that hepatic BMP2 levels were significantly more elevated in NASH than in NAFL patients (p=0.0047). Notably, a significant positive correlation in all study population was observed (p<0.0001, Figure 1C) between the hepatic mRNA levels of BMP2 and CD36, a fatty acid transporter which contributes to liver fat accumulation in patients with NAFLD ^23^.

**Figure 1.**
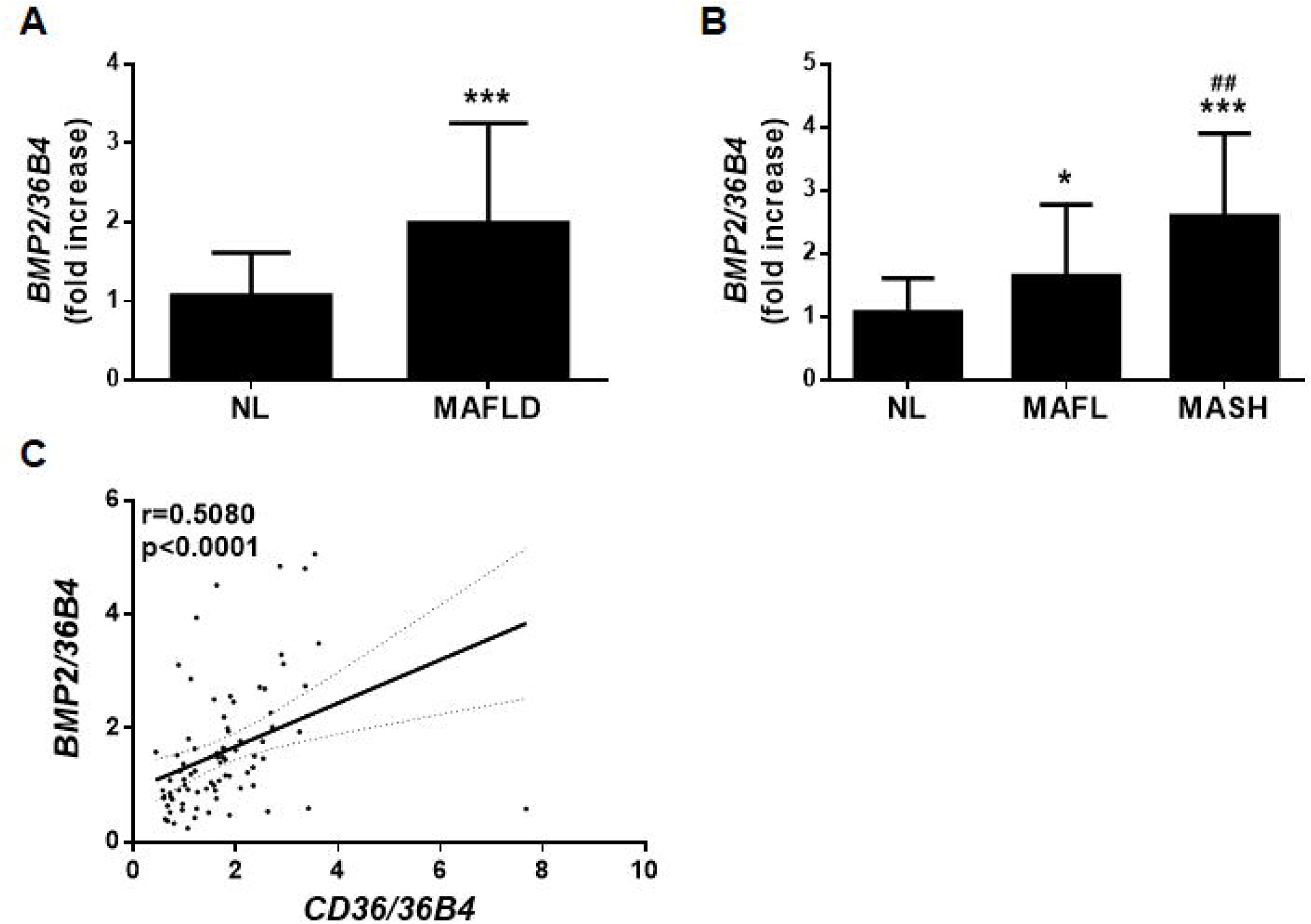
Expression of BMP2 is increased within the liver of NAFLD patients. **A and B**. Hepatic *BMP2* mRNA levels determined by RT-qPCR and normalized to *36B4* gene expression. Data are expressed as fold increase relative to control condition (1) and presented as mean ± SD. **C**. Correlation in the study population of matched mRNA expression levels (*BMP2* and *CD36*). <u>Study population</u>: Normal liver (NL) individuals (n=75), NAFLD patients (n=115: 56 NAFL and 59 NASH). *p<0.05 and ***p<0.005, NAFLD, NAFL or NASH vs. NL; ^##^p<0.01, NASH *vs*. NAFL.

### Serum levels of BMP2 are elevated in NAFLD patients

Next, we measured the concentration of BMP2 in serum samples from all NAFLD patients and NL subjects included in our study. Mean circulating BMP2 levels were significantly higher in NAFLD patients than in NL subjects (p=0.0033; Figure 2A). Notably, serum BMP2 concentrations were more elevated in both NAFL and NASH patients compared to NL subjects (p=0.0552 and p=0.0025, respectively; Figure 2B). Interestingly, no significant differences were found between patients with NAFL and NASH. In addition, a significant and progressive increase in serum BMP2 levels was found in NAFLD patients in relation to the histological grade of steatosis (p=0.0032, Figure 2C) and NAFLD activity score (p=0.0081, Figure 2D). However, circulating BMP2 levels did not correlate with either BMI (p=0.9162, Figure 2E) or HOMA-IR (p=0.4420, Figure 2F), indicating that serum BMP2 levels in patients with NAFLD are not influenced by adiposity or insulin resistant state.

**Figure 2.**
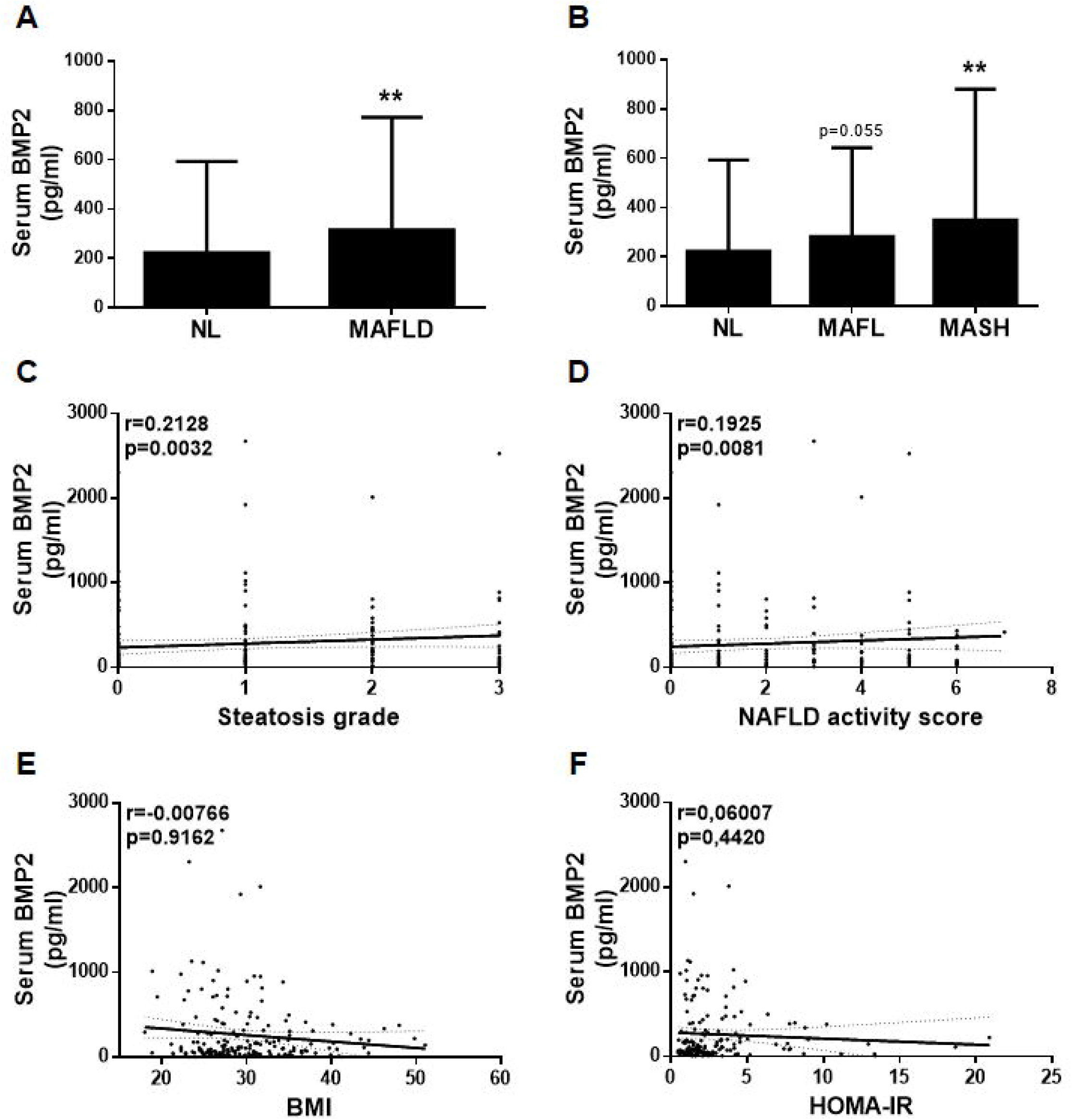
Serum levels of BMP2 are increased in NAFLD patients. **A and B**. Serum levels of BMP2 determined by ELISA. Data are expressed as pg/ml and presented as mean ± SD. **C**. Correlation in the study population of matched serum BMP2 levels with the steatosis grade. **D**. Correlation in the study population of matched serum BMP2 levels with the NAFLD activity score. **E**. Correlation in the study population of matched serum BMP2 levels with the BMI. **F**. Correlation in the study population of matched serum BMP2 levels with the HOMA-IR. <u>Study population</u>: Normal liver (NL) individuals (n=75), NAFLD patients (n=115: 56 NAFL and 59 NASH). **p<0.01, NAFLD, or NASH *vs*. NL.

### BMP2 expression in human hepatocytes

In order to clarify which liver cells can be the cellular source of BMP2, we performed some *in vitro* experiments using human hepatocytes (Huh7 cells) treated with different doses of palmitic acid (PA) for 16 hours, a known protocol for inducing intracellular lipid accumulation^24^. After the exposure to PA, results obtained revealed an elevation of intracellular lipid content (Figure 3A) in parallel to an increase of mRNA levels of BMP2 in Huh7 hepatocytes (Figure 3B). Moreover, an increase of supernatant BMP2 concentration was also observed upon PA treatment (Figure 3C).

**Figure 3.**
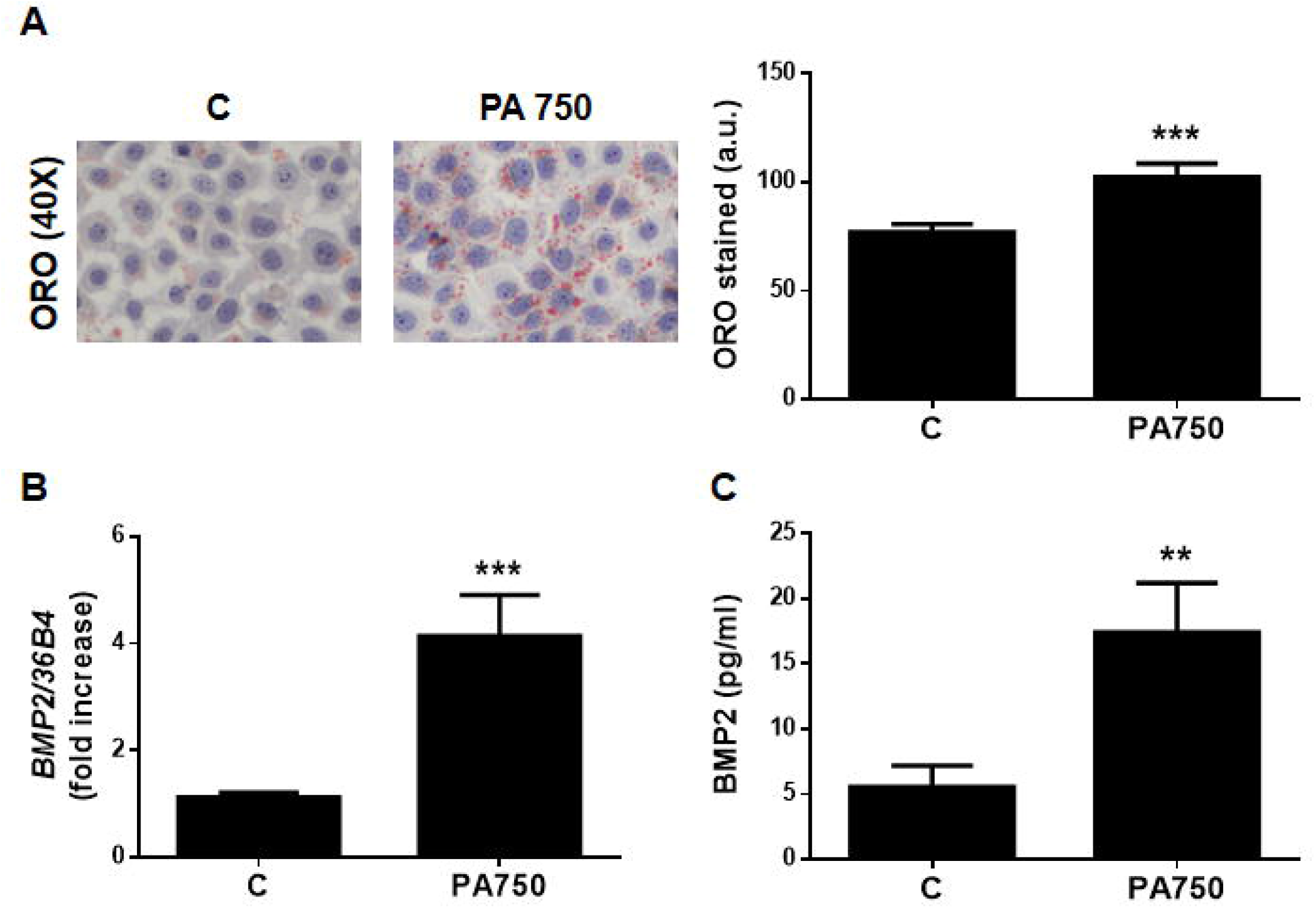
Palmitic acid overload upregulates mRNA levels of BMP2 in Huh7 hepatocytes. **A**. Representative images of ORO staining and its quantification. **B**. Hepatic *BMP2* mRNA levels were determined by RT-qPCR and normalized to *36B4* gene expression. Data are expressed as fold increase and presented as mean ± SEM relative to control condition (1). **C**. BMP2 levels in the cell supernatant determined by ELISA. Data are expressed as pg/ml and presented as mean ± SEM. <u>Experimental conditions:</u> Huh7 cells treated with 750 mM palmitic acid (PA) for 16 h (n=3 independent experiments performed by duplicate). **p<0.01 and ***p<0.005, PA *vs*. C.

### Circulating BMP2 is an independent predictor of NASH and the algorithm SAN is able to discriminate NASH

Since serum BMP2 levels were significantly higher in NASH than in NL subjects, we performed logistic regression analysis in the entire study population in order to determine whether serum BMP2 levels are associated with NASH. In order to find the best sensibility and specificity threshold for NASH diagnosis, the best cut-off value for serum BMP2 concentrations was established by using the Youden index at 49.21 pg/mL. Circulating BMP2 was identified as an independent predictor of NASH in univariate analysis (OR [95% CI], 3.5 [1.47-8.37], p=0.005; Table 2). Multivariate analysis, adjusted for potential confounders (age, gender, BMI) and variables clinically relevant to NAFLD (glucose and gamma-glutamyltransferase -GGT-) showed that serum BMP2 concentrations persisted significantly associated with NASH (OR [95% CI], 5.07 [1.55-16.6], p=0.007; Table 2). Thus, we performed ROC curve analysis to determine the accuracy of the algorithm termed SAN (Screening Algorithm for NASH), based in a formula derived from the previously mentioned logistic regression model (Figure 4), which revealed a good accuracy to discriminate NASH (AUROC, 0.886). Next, 0.1 unit intervals of SAN were analyzed to establish best cut off points (Table 3). A SAN ≤ 0.2 can be used to exclude NASH (SN = 0.84; NPV=0.909) and a SAN ≥ 0.6 to rule it in (SP = 0.96; PPV=0.86).

**Table 2.** Univariate and multivariate analysis of the independent variables associated with NASH in the study population.

| Independent variables | Univariate analysis |  |  | Multivariate analysis |  |  |
| --- | --- | --- | --- | --- | --- | --- |
|  | OR | 95% CI | p value | OR | 95% CI | p value |
| BMP2 (pg/ml)<br>Cut-off = 49.21 | 3.5 | [1.47-8.37] | 0.005 | 5.07 | [1.55-16.6] | 0.007 |
| Gender (female/male) | 2.77 | [1.43-5.36] | 0.002 | 2.96 | [1.22-7.14] | 0.016 |
| Age (years) | 1.04 | [1.01-1.06] | 0.004 | 1.03 | [0.99-1.06] | 0.06 |
| BMI (kg/m<sup>2</sup>) | 1.22 | [1.14-1.31] | <0.001 | 1.28 | [1.16-1.41] | <0.001 |
| Glucose (mg/dl) | 1.03 | [1.01-1.04] | <0.001 | 1.02 | [1-1.04] | 0.018 |
| GGT (IU/L) | 1.01 | [1.01-1.02] | 0.001 | 1.01 | [1-1.02] | 0.016 |

OR, odds ratio; CI, confidence interval; BMI, body mass index; GGT, gamma-glutamyltransferase.

**Table 3.**
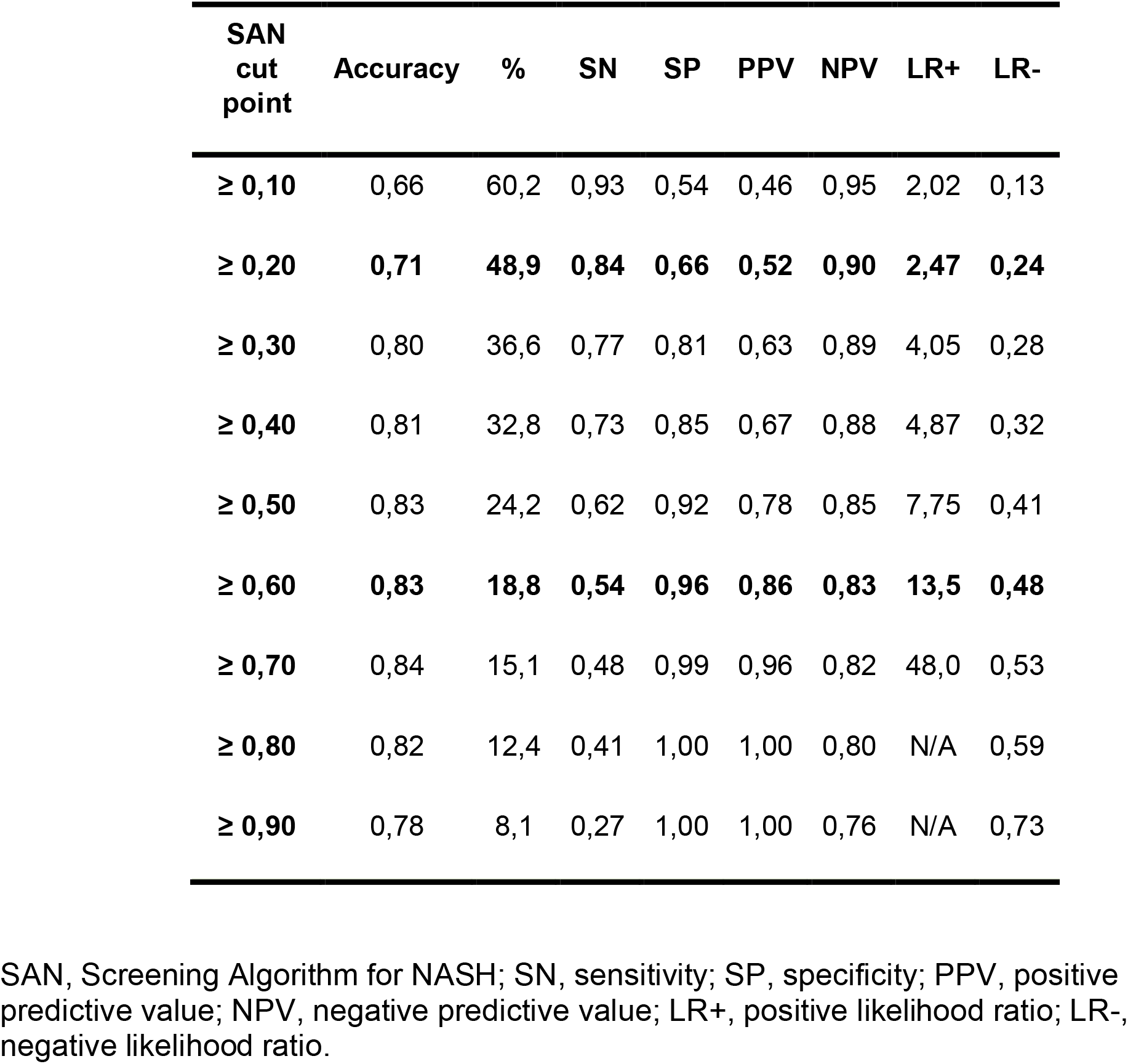
Diagnostic accuracy of SAN.

**Figure 4.**
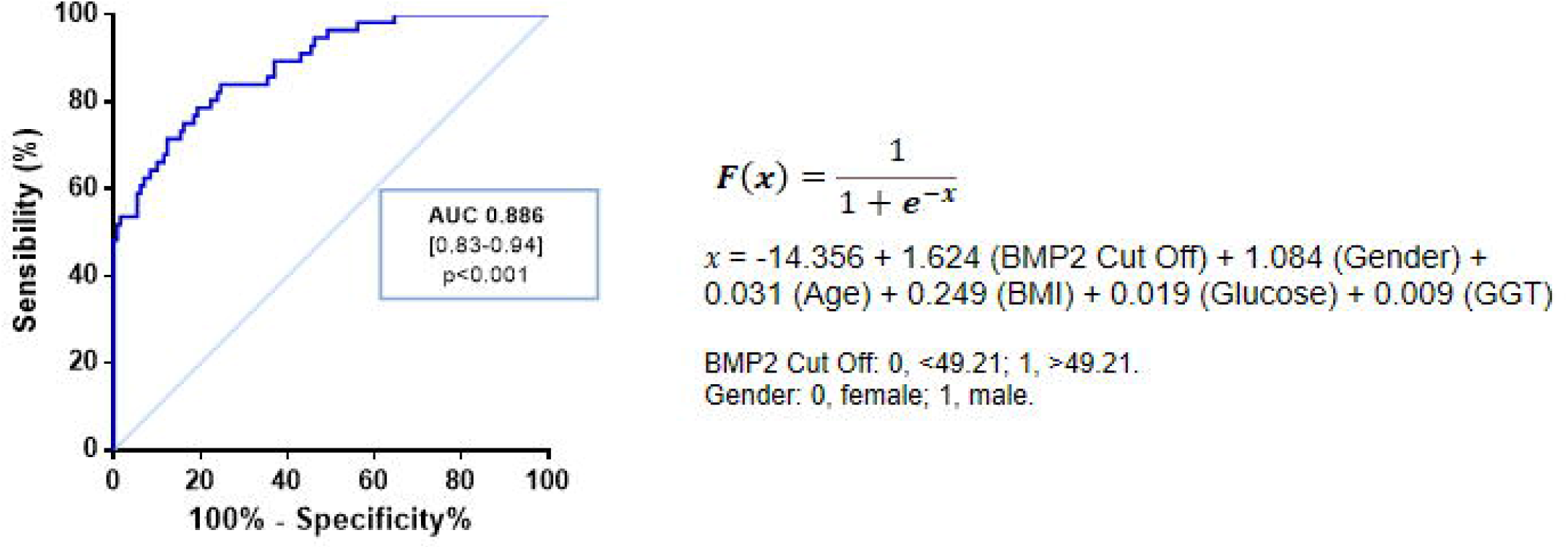
Receiver operating characteristic (ROC) curve showing diagnostic accuracy of SAN algorithm for detecting NASH. AUROC, area under the ROC curve; BMI, body mass index; GGT, gamma-glutamyltransferase.

## DISCUSSION

Nonalcoholic fatty liver disease (NAFLD), is the major cause of chronic liver disease worldwide, with a diverse histopathological spectrum ranging from simple steatosis without significant inflammation, also termed fatty liver (NAFL), to steatohepatitis (NASH) with varying stages of fibrosis and, ultimately, cirrhosis and hepatocellular carcinoma ^25^.

Given that NAFLD is usually asymptomatic at the time of diagnosis, the determination of the lipid content within the liver is an important clinical challenge in terms of early identification to allow appropriate therapeutic interventions in order to prevent the progression of liver disease to more advanced fibrotic states or, even, to the end-stage liver disease. In the last decade, there has been an explosion in the discovery of biomarkers for NAFLD diagnosis and/or prognosis. Considering the wide range of their potential applications in clinical practice, the growing interest of the academic and industrial community in biomarkers is not surprising. Indeed, lack of awareness about risk factors for NAFLD and its progression, combined with insufficient or unreliable screening and surveillance modalities, may contribute to a delay in diagnosis and may explain why many patients are diagnosed in the later stages of the disease ^7^.

A novel finding of the present study is that BMP2 expression is increased in the liver from NAFLD patients as well as that serum BMP2 levels were also higher in these patients than in subjects with histologically normal liver, positioning this protein as a new molecular target linked to NAFLD. One limitation of this study is the limited number of individuals taking into account that women were predominant in all the histological groups studied, especially within patients with histological NL, and NAFLD patients were significantly older than NL subjects. In fact, BMP2 expression might be affected by age and sex. However, no correlation was found between circulating BMP2 levels and age (*r=*-0.05484 *p=*0.4524), and there were no differences when separating the population by sex (*p=* 0,2035). In addition, we also shown herein that human hepatocytes are able to produce and secrete BMP2 upon stimuli with PA mimicking steatotic conditions ^24^, indicating that BMP2 might be involved in hepatosteatosis development, not only in the fibrogenic process as some studies have previously reported _26,27_.

Interestingly, there are only two studies so far showing that the expression of some BMPs, specifically BMP6 and BMP8B, is increased in the liver of NAFLD patients ^13,28^. These studies, along with our findings, indicate that BMPs might play a role in NAFLD pathophysiology. In this regard, it has been demonstrated that the pharmacological inhibition of BMP signaling reduced diet-induced hepatic steatosis in obese mice by targeting the production of triglycerides in the liver ^18^; but also, this study has also shown that this effect is specific for the inhibition of BMP type I receptor ALK3, proposing a role of BMP2 and BMP4 in the establishment of NAFLD ^18^.

Going further, our study is the first which reports circulating BMP2 concentrations in NAFLD patients. Other authors have previously reported that serum levels of distinct BMPs varied widely in the context of other metabolic disorders. For instance, elevated serum BMP4 levels have been correlated with carotid atherosclerosis in T2D patients ^29^. Conversely, various studies have associated BMP9 with glucose and lipid metabolism because a decrease in the circulating concentrations of BMP9 have been observed in patients who displayed features of metabolic syndrome such as insulin resistance, T2D or arterial hypertension ^30-32^. BMP9 has also been related to other diseases, such as portopulmonary hypertension, with lowered levels of this protein acting as a potential risk factor ^33^. Regarding liver diseases, while BMP7 has been proposed as a marker for hepatic carcinogenesis because its serum levels were found to be decreased in patients with liver cancer ^34^, other authors have reported that circulating BMP7 levels are augmented in patients with viral chronic liver diseases ^35^. Considering BMP2 reports, some studies have associated increased levels of this BMP with coronary artery disease in diabetic patients ^36^. Moreover, an elevation of serum BMP2 concentrations was observed in overweight and obese middle-aged and elderly women ^37^, whereas in the present study, a correlation between circulating levels of BMP2 and BMI was not observed in agreement with results recently reported by Lopes *et al* ^38^. Nevertheless, further investigations are warranted to elucidate the role of BMP2 regulating glucose and lipid homeostasis and its impact in the pathogenesis of metabolic diseases such as obesity, diabetes and NAFLD.

One of the most striking findings of the present study is that serum BMP2 levels were significantly associated with biopsy-proven NASH in multivariate logistic regression model, suggesting that BMP2 might have utility as biomarker to screen NASH in different clinical settings. Using serum BMP2 levels combined with age, gender, BMI, glucose and GGT, an algorithm termed SAN (Screening Algorithm for NASH) was derived from a logistic regression model which discriminated NASH patients from subjects with histologically normal liver or NAFL. This is a breakthrough because there are no validated non-invasive tools for predicting NASH so far, the most clinically relevant form of NAFLD which can only be diagnosed by liver biopsy. In our population, a value of SAN ≥ 0.6 indicated a high risk for biopsy-proven NASH with a good accuracy (AUROC, 0.86; SP, 0.96) showing that SAN may be useful to screen populations at risk for NAFLD.

In conclusion, this proof-of-concept study provides the first scientific evidence that hepatic and serum levels of BMP2 are abnormally elevated in NAFLD patients, indicating that BMP2 is a new molecular target linked to human NAFLD. In addition, we describe a simple and efficient algorithm termed SAN which was able to discriminate NASH with a good accuracy in our study population. Further studies, however, in distinct and larger cohorts of NAFLD patients are needed to validate the potential utility of SAN as a non-invasive algorithm for NAFLD evaluation.

## Data Availability

All data produced in the present study are available upon reasonable request to the authors

## Acknowledgements

We thankfully acknowledge Elvira del Pozo for helpful clinical assistance. This work was supported by grants GLD18/00151 from Gilead Science and PI20/00837 from Instituto de Salud Carlos III (ISCIII, Spain) and Fondo Europeo para el Desarrollo Regional (FEDER) and CIBEREHD (ISCIII) to CGM, and contracts CP14/00181 and CP19/00032, and grants PI16/00853 and PI19/00123 from ISCIII/FEDER (Spain) and CIBERDEM (ISCIII) to AGR.

## Conflict of Interest

All authors have declared that no competing interest exists.

## Notes

**Funding support.** This work was supported by grants GLD18/00151 from Gilead Science and PI20/00837 from Instituto de Salud Carlos III (ISCIII, Spain) and Fondo Europeo para el Desarrollo Regional (FEDER) and CIBEREHD (ISCIII) to CGM, and contracts CP14/00181 and CP19/00032, and grants PI16/00853 and PI19/00123 from ISCIII/FEDER (Spain) and CIBERDEM (ISCIII) to AGR.

### Competing Interest Statement

The authors have declared no competing interest.

### Author Declarations

The Human Ethics Committee of the Hospital Universitario Santa Cristina (reference, PI-688A) and Hospital Universitario Virgen del Rocio (reference, C.I. 0359-N-15) approved the study procedures, and all participants signed an informed written consent before inclusion in the study.

